# SARS-CoV-2 quarantine mandated by contact tracing: burden and infection rate among close contacts in Zurich, Switzerland, 2020-2021

**DOI:** 10.1101/2023.09.03.23294798

**Authors:** Hélène E. Aschmann, Anja Domenghino, Ruedi Jung, Tala Ballouz, Dominik Menges, Jan Fehr, Milo A. Puhan

## Abstract

**Objectives:** Before vaccines and effective treatments were available, quarantine of close contacts was important to limit the spread of SARS-CoV-2. To evaluate potential benefits and harms of quarantine, we aimed to estimate infection rates and describe experiences and mental health among persons in mandated quarantine during the early SARS-CoV-2 pandemic.

**Methods:** We invited adults in mandated quarantine after an exposure to SARS-CoV-2 identified through contact tracing of the Canton of Zurich, Switzerland, between August 2020 and January 2021. Participants completed two questionnaires and received up to two SARS-CoV-2 polymerase chain reaction tests, during and at the end of quarantine.

**Results:** Among 395 participants, quarantine duration ranged from 2 to 20 days. By day 11 since the last contact, 11.1% [95% CI 8.4%–14.7%] were infected with SARS-CoV-2. The proportion of participants with symptoms of depression doubled from 9.3% before quarantine to 18.9% during quarantine, and 12.1% reported quarantine was very or extremely difficult.

**Conclusions:** Although quarantine was only moderately burdensome for most participants, some experienced significant difficulties and burden. Policymakers need to balance infection control with potential harms placed on individuals.

## Introduction

Authorities mandating quarantine of close contacts of persons infected with SARS-CoV-2 need to balance quarantine’s efficacy in limiting viral spread against its impact on individuals, society, and economy. The balance of quarantine’s benefits and harms should not only be counted with mortality and morbidity, but should also consider protection from financial, social, and psychological harm on quarantined persons [1]. Quarantine was especially important at the beginning of the SARS-CoV-2 pandemic when vaccines and effective medical treatment were not available. Currently, only few countries still impose quarantine and in Switzerland, quarantine mandates were lifted in April 2022. But with new variants and waning vaccine effectiveness, quarantine remains an option according to the World Health Organization [2].

Quarantine is associated with poor psychological outcomes, in particular after a duration of one week or longer [3,4]. Individuals in quarantine or isolation have 2-3 times increased odds for depressive, anxiety or stress-related disorders [4]. Furthermore, low adherence (42% in the UK [5], 28% in Norway [6]) limited its efficacy. While at the beginning of the pandemic, some called for quarantine as long as 21 days based on incubation time [7–9], testing soon allowed to reduce quarantine duration [10–15]. Later some persons were exempted from quarantine due to prior infections or vaccination [2].

To evaluate or inform policy makers, a detailed evaluation of the benefits and harms of quarantine measures is warranted. Our aims were (1) to estimate infection rates among close contacts in mandated quarantine in the Canton of Zurich, Switzerland, during the early SARS-CoV-2 pandemic (i.e., the secondary attack rate), and (2) to assess mental health, difficulties and worries during quarantine as well as adherence, and motivation to adhere to quarantine.

## Methods

### Study Population

This analysis is based on the Zurich SARS-CoV-2 Cohort, a prospective, observational, population-based study of an age-stratified random sample of 1106 SARS-CoV-2 positive individuals and their close contacts based on contact tracing in the Canton of Zurich, Switzerland. All participants provided electronic consent. The study was prospectively registered (ISRCTN14990068) and approved by the ethics committee of the Canton of Zurich (BASEC 2020-01739).

More detailed information about the study enrolment procedures are reported elsewhere [16]. In short, close contacts were identified through contact tracing and randomly sampled in clusters based on their index case (the person with a positive SARS-CoV-2 test in the cohort with whom a potentially infectious contact occurred). We recruited close contacts from August 7, 2020, to January 15, 2021. Eligible persons included adults (aged ≥18), fluent in German, resident in the canton of Zurich, and consenting to participate. Persons were not eligible if they declined to be re-contacted for study purposes or if the index case was not recorded (e.g., the contact occurred in another country).

### Data collection

Close contacts were invited to complete a baseline questionnaire upon enrolment. A second questionnaire was sent on the second to last day of quarantine. Moreover, they were invited to receive a polymerase chain reaction (PCR) test for SARS-CoV-2 at the beginning of quarantine (preferably on day 5 or 6) and at the end of quarantine. Close contacts testing positive were invited to participate in a separate arm of the study instead. We reported mental health outcomes in persons who tested positive separately [17].

The questionnaires included the German version of short form of the Depression, Anxiety, and Stress Scale (DASS-21) [18], and a subset of questions from the COVID-19 Pandemic Mental Health Questionnaire [19]. We asked how well informed and prepared participants felt, how difficult quarantine measures were in general to follow, and how difficult the specific rules published by the Swiss Federal Office of Public Health were [20]. Potential worries, reasons for motivation and positive experiences were piloted with lay persons and we added open text comments to allow participants to add others.

To compare participants with non-participants, aggregated data were analysed based on data from contact tracing. These data were recorded in Microsoft Office Excel forms and in the Surveillance Outbreak Response Management and Analysis System (SORMAS) [21] a web application developed for contact tracing.

### Sample size estimation

The sample size calculation was based on the proportion of close contacts with a positive PCR test after day 5 of quarantine. We estimated 5% would test positive until day 5, and 3% with an initial negative test would test positive by the end of quarantine. To show a difference of 1% at a significance level of 0.05, a sample size of 294 was needed. However, the sample size was increased because not all participants agreed to get tested. Recruitment was stopped when the targeted sample size was reached for SARS-CoV-2 positive individuals of our cohort study, as the study had several aims.

### Statistical analysis

We modelled the cumulative probability over time of converting during the mandated quarantine for all invited close contacts. This cumulative probability was derived by modelling the proportion of persons who remained without a positive test using a survival analysis with the tram package (version 0.6-4) in R [22]. For each positive test, we assumed the conversion to occur earliest on the day after the last negative test or, in the absence of a test, on the day of last contact, and latest on the day the sample for the positive test was taken (interval censoring) [23,24]. For close contacts without a positive test, the observations were censored after the last negative test. Because our recruitment in the close contact sub-study did not include contacts who converted before they consented to participate, we also included data from non-participants in the model, to account for contacts who converted early. For non-participants, we obtained the sampling day of positive tests from contact tracing data. We assumed a similar frequency of negative tests as for participants, except for the tests on day 10 and 11 of quarantine. Since testing at the end of quarantine was a study-specific procedure, and not mandated by the Department of Health, we assumed non-participants had only as many negative tests on day 10 and 11 as participants on day 9.

Descriptive analyses were performed for all other outcomes. For DASS-21, missing data were imputed for each subscale if two or less answers were missing using the mean of the remaining answers. We explored associations of (1) overall motivation, (2) feeling prepared, (3) difficulty of quarantine measures overall, and (4) having more time to relax during quarantine with baseline characteristics based on a priori hypotheses and hypotheses generated based on participants’ comments. We included age and sex in all regressions. All outcomes were elicited as five-point Likert-type scales, and we used logistic ordinal regression to analyse associations [25].

Informed consent, recruitment and survey data were collected and managed using REDCap electronic data capture tools [26,27]. For participants who preferred not to complete the survey online, written consent was sought via letters and questionnaires were completed via phone. All analyses were performed using R version 4.1.1 [28].

## Results

### Study Population

Of 21 316 records of close contacts, 10 110 were considered eligible during the initial screening (Figure 1). We invited 1484 persons (and 8 duplicates). Of these, 129 were not eligible, and 526 had finished quarantine or converted to a positive case. Of the remaining 829 persons, 395 consented to participate, corresponding to a response rate of 48%.

**Figure 1:**
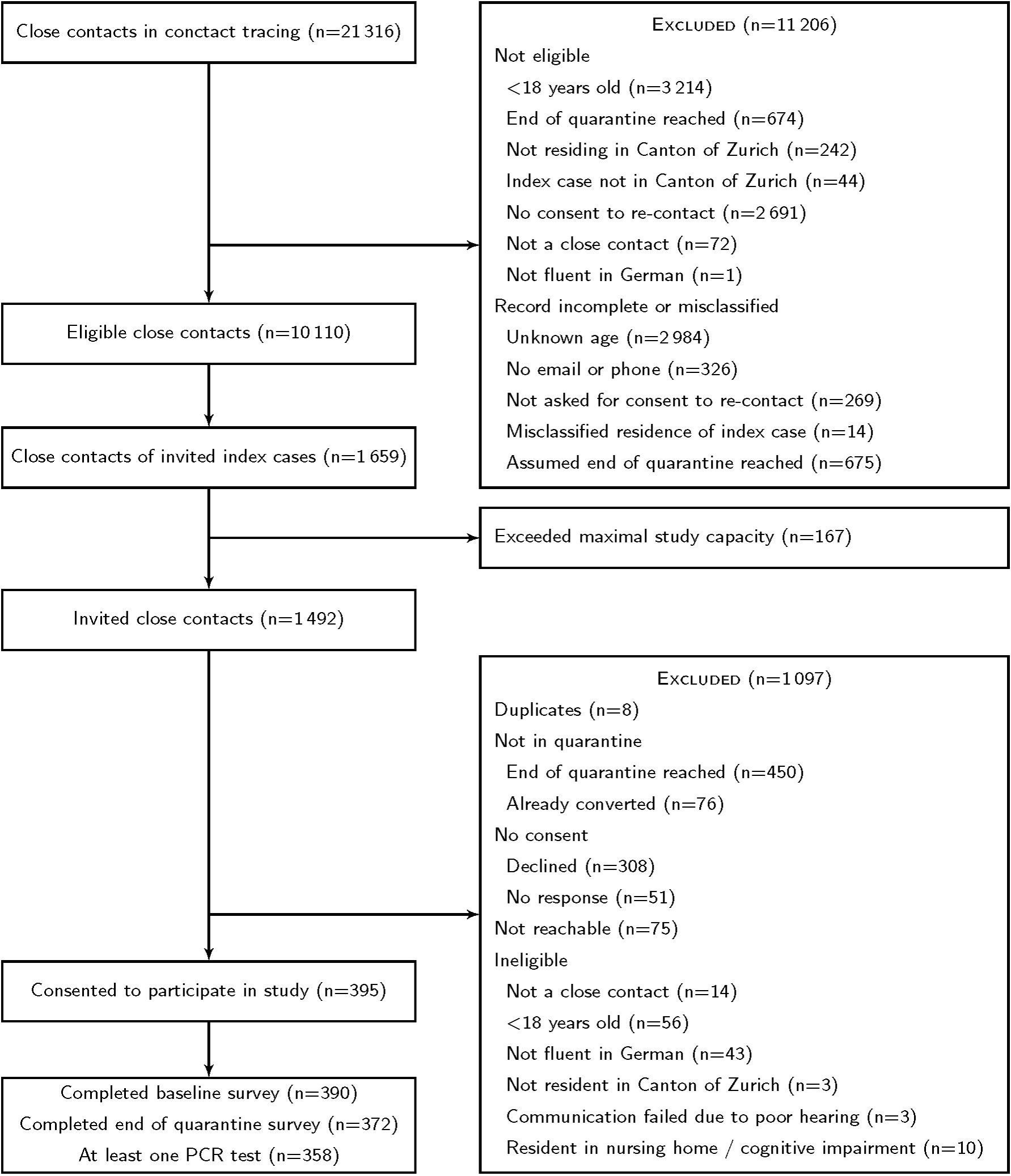
Participant flow through recruitment. Recruitment was performed in two stages. Potential participants had to complete an online form to be contacted by the study team or give oral consent to have the online form completed for them.

We compared baseline characteristics between 395 participants and 1018 non-participants. This analysis did not consider 79 persons who should not have been invited (8 duplicates, 56 underage, 14 not close contacts, 1 already converted before we invited them). Participants and non-participants were similar in sex and duration of quarantine (Table S1). Age was missing for 36% of non-participants, so a direct comparison is difficult. However, participants were likely younger than non-participants because their index cases (persons with a positive SARS-CoV-2 test) were younger (Table S1), and the age of index cases and close contacts was correlated (Table S2).

Almost all participants (94%) performed the mandated quarantine at home (Table 1). Some participants (22%) had children in their household. Many were household contacts, meaning they were living in the same household as an individual with a SARS-CoV-2 infection (35%). Most participants were employed (70%). Of those who worked, most could work at home (47%) or at least partially (24%). Some self-quarantined before receiving the official mandate because they knew of a contact with an infected person (38%). Among household contacts, 47% (64/135) self-quarantined, while 37% (50/135) answered that their reason to initiate quarantine was the official mandate.

**Table 1:**
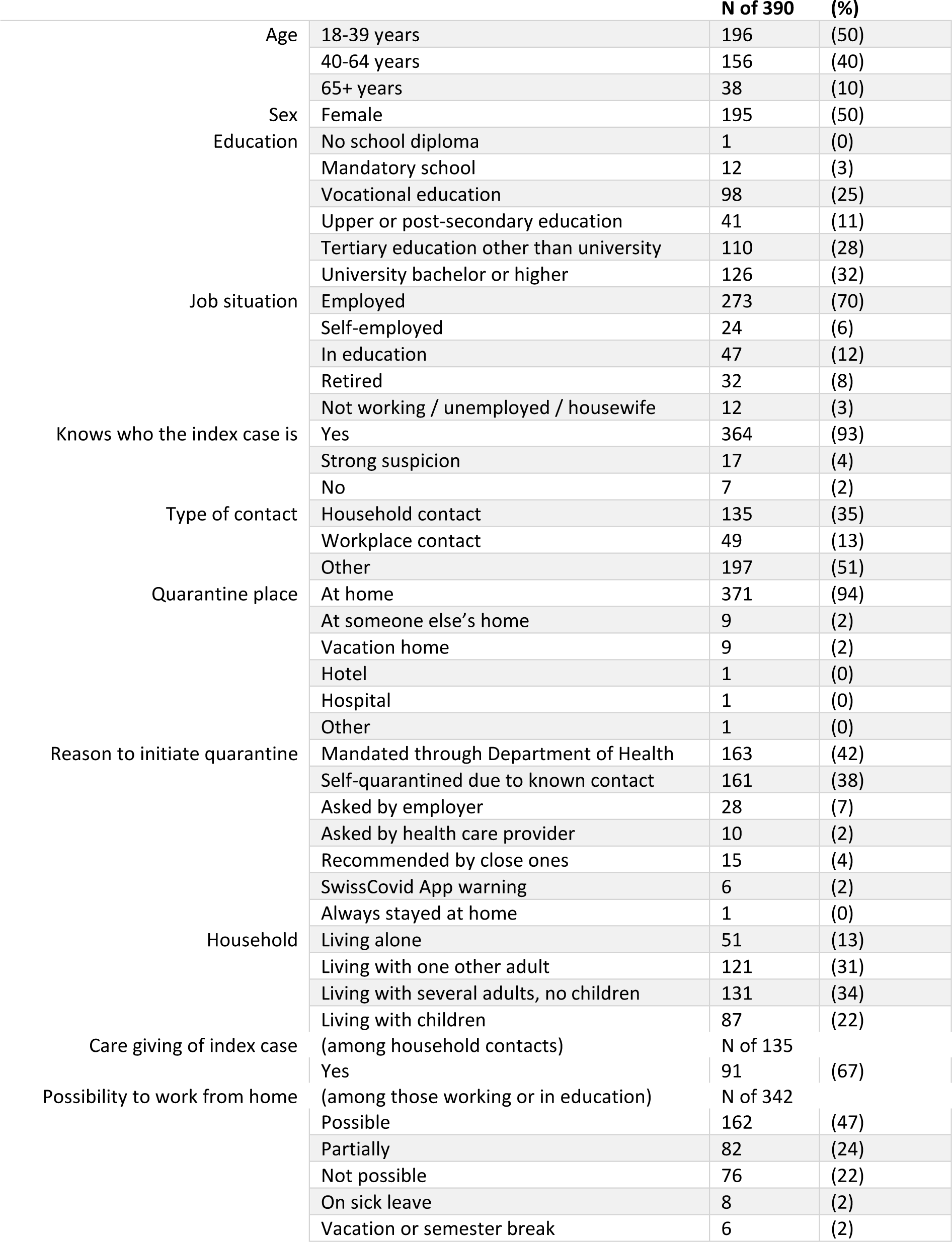
Baseline characteristics of participants and information relevant to their quarantine.

### PCR test results during quarantine

At least one PCR test was performed in 358 of 395 participants during their quarantine. Our analysis of the secondary attack rate considered a total of 1404 invited adult close contacts in whom we could have noted a conversion (Figures S1 and S2). In a survival analysis that considers when persons were tested (positive and negative), the estimated cumulative probability for a conversion by day 11 was 11.1% [95% CI: 8.4–14.7%] (Figure 2A). Most conversions occurred within 5 days after the last potentially infectious contact: the estimated cumulative probability reached 9.1% [95% CI: 6.6– 12.7%] by day 5. The mean follow-up time up to the last test result was 7.4 days.

**Figure 2:**
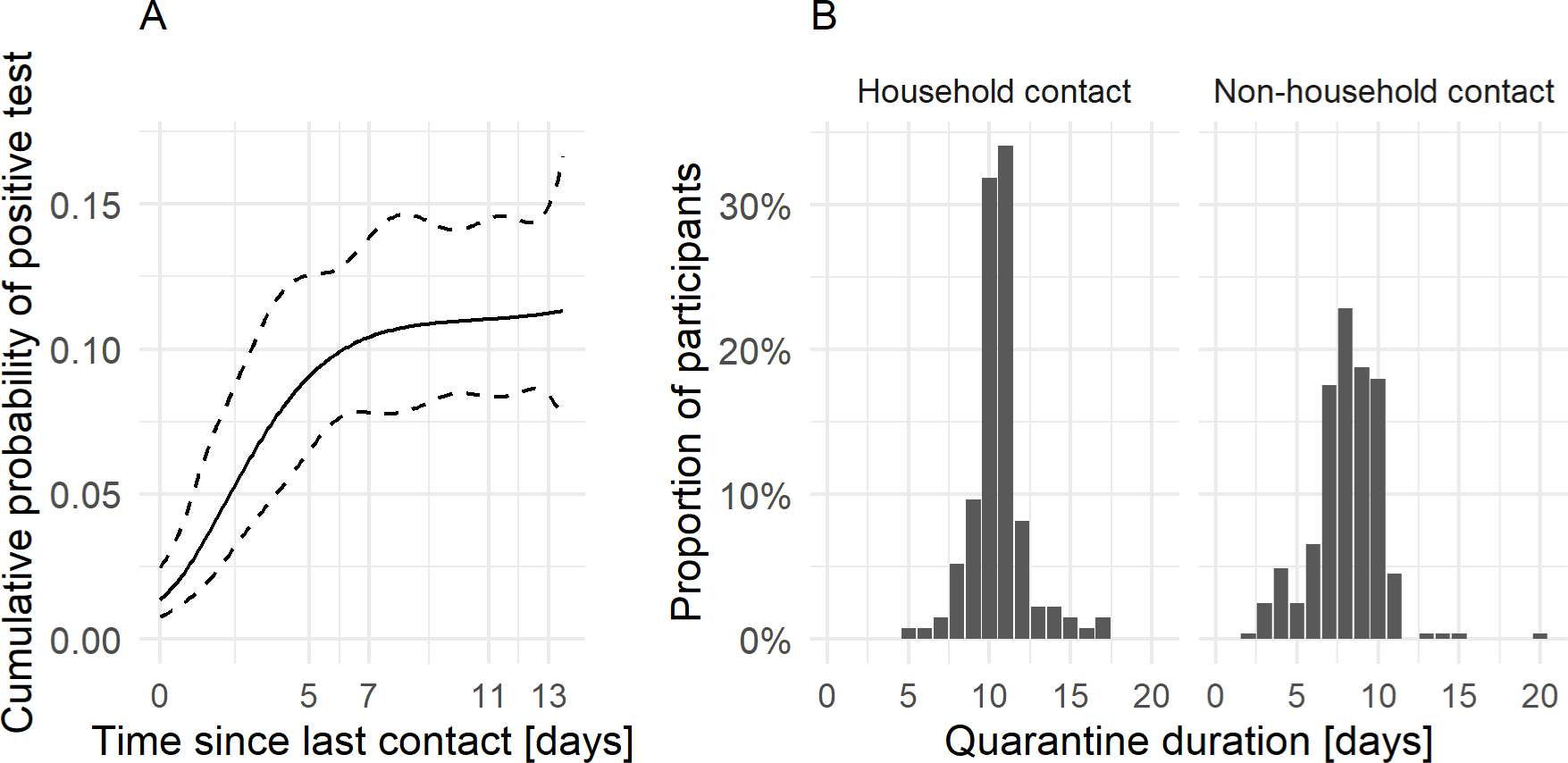
Timing of conversions during quarantine and duration of quarantine A) Cumulative probability of conversion from close contact to case with positive test (i.e., secondary attack rate), from the day of last contact with the index case. Most conversions occur within the first 5 days since last contact. Conversion on day 0 means that the sample for the PCR test was taken on the day of last contact. The person was reported as a close contact and most likely received the positive test result one day later. B) Duration of quarantine for household contacts and non-household contacts. This includes potential self-quarantine before the mandated quarantine and extensions due to other household members in quarantine converting to cases.

### Quarantine duration

In theory, household contacts should always have had a quarantine duration of at least 10 or 11 days (quarantine duration was decreased from 11 to 10 days in November 2020). However, 24 of 135 household contacts (17.8%) reported durations of 5 to 9 days (Figure 2B). A regular duration of 10 or 11 days was reported by 89 (65.9%) and an extended duration by 22 (16.3%) household contacts. Non-household contacts declared shorter durations of quarantine, with a median of 8 days and ranging from 2 to 20 days. Among all contacts, quarantine was extended in 29 (7.4%) persons; this occurred when persons spent their quarantine together and one of them converted during that time.

### Motivation to adhere to quarantine measures

Most participants reported that they were motivated to adhere to quarantine measures overall: 338 (86.7%) answered that they were motivated, very motivated, or extremely motivated (Figure 3). Participants were most motivated to get tested when symptoms appeared (360, 92.3%). However, less participants were motivated not to leave their home for 10 days (275, 70.5%), not to meet family and friends (269, 69.0%), to follow hygiene rules (220, 56.4% among those not living alone), and to stay at home longer if necessary (198, 50.8%) (e.g., if they or others in their household developed symptoms).

**Figure 3:**
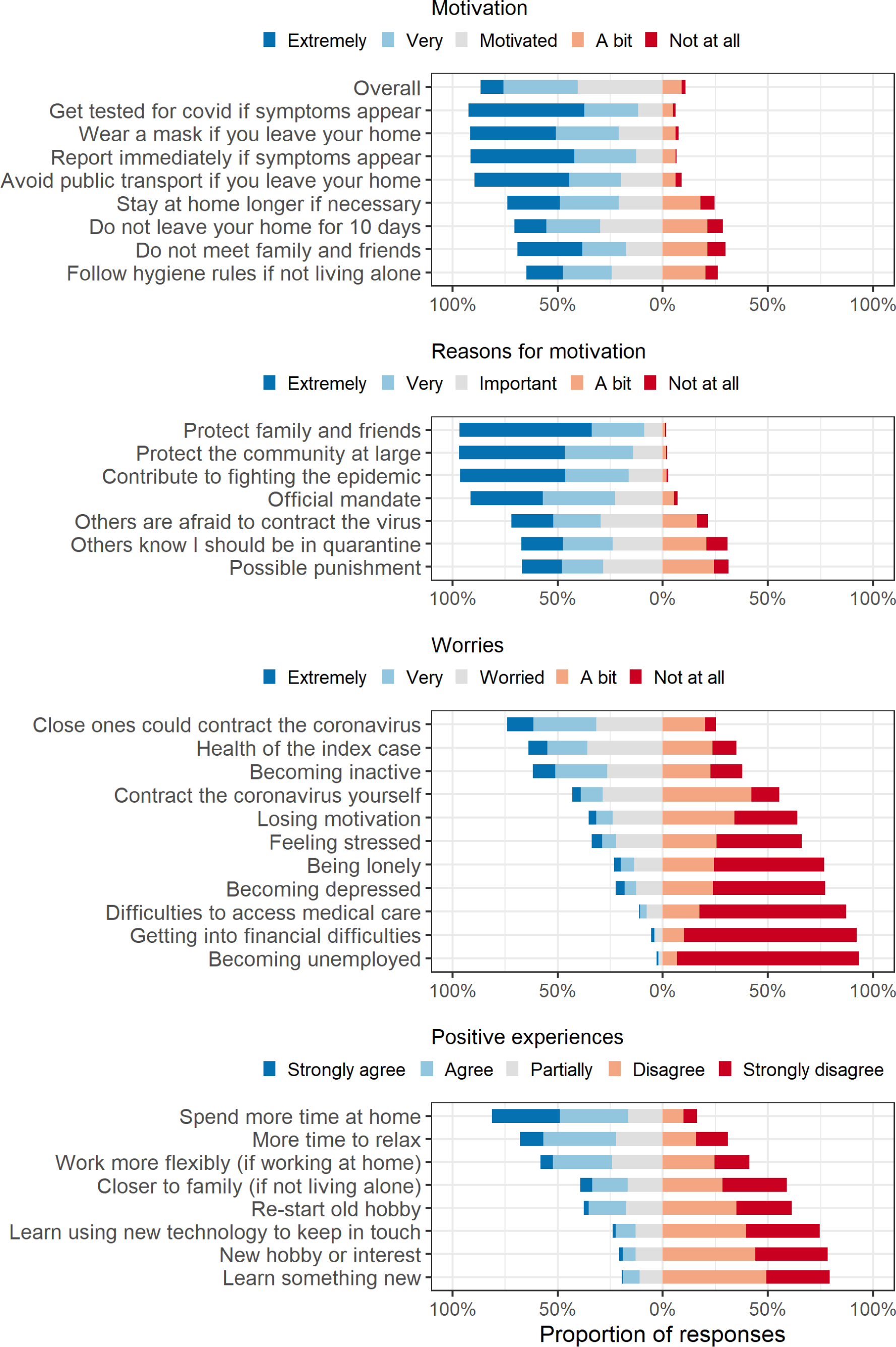
Participants’ responses regarding motivation for quarantine measures, reasons for motivations, worries, and positive experiences during quarantine.

The most important motivations to stay in quarantine were to protect family and friends, protect the community at large, and contribute to fighting the pandemic, followed by the official mandate from authorities (Figure 3). Less participants considered others knowing they should be in quarantine, others being afraid to contract the virus, or possible punishments as important reasons.

### Worries during quarantine

Participants worried most that their loved ones could contract the coronavirus: 289 (74.1%) were worried, very, or extremely worried (Figure 3). Most worried about the health of the person infected with SARS-CoV-2 (i.e., their index case) (63.8%) and becoming inactive (61.8%). Many worried about contracting the virus themselves (43.1%). Some participants worried about becoming depressed (22.3%).

Worries were associated with the overall motivation (Table 2). Participants who worried about contracting the virus themselves had 2.08 [95% CI: 1.41–3.09] times higher odds of reporting higher levels of overall motivation. By contrast, worries about becoming depressed were associated with lower motivation (0.45 [95% CI: 0.28–0.72] times smaller odds for higher levels of motivation).

**Table 2:** Associations of baseline participant characteristics with motivation, feeling prepared, difficulty with quarantine measures, time to relax (all ordinal logistic regressions where the outcome was a five-point Likert-type scale), and change in depression score (linear regression). *Statistically significant at 0.05 significance level, not corrected for multiple testing.

| **Outcome: Higher overall motivation to comply with quarantine measures** |  | **OR [95% CI]** |
| --- | --- | --- |
| Age (per 10 years, reference age 18) |  | 1.01 [0.90–1.14] |
| Sex (male vs female) |  | 0.96 [0.66–1.40] |
| Worried about getting sick with COVID-19 (own health) (worried, very worried, or extremely worried vs little or not at all worried) |  | 2.08 [1.41–3.09]* |
| Worried about becoming depressed (worried, very worried, or extremely worried vs little or not at all worried) |  | 0.45 [0.28–0.72]* |
| **Outcome: Feeling better prepared** |  |  |
| Age (per 10 years, reference age 18) |  | 1.19 [1.05–1.34]* |
| Sex (male vs female) |  | 0.91 [0.62–1.33] |
| Well or very well informed (vs neutral, poor, or very poor) |  | 5.27 [3.03–9.32]* |
| Worried about becoming depressed (worried, very worried, or extremely worried vs little or not at all worried) |  | 0.37 [0.23–0.58]* |
| Worried about getting sick with COVID-19 (own health) (worried, very worried, or extremely worried vs little or not at all worried) |  | 0.96 [0.65–1.42] |
| **Outcome: Overall difficulty with quarantine measures** |  |  |
| Age (per 10 years, reference age 18) |  | 0.84 [0.75–0.95]* |
| Sex (male vs female) |  | 0.50 [0.34–0.72]* |
| Working (possible or partially possible vs not possible) |  | 1.00 [0.67–1.47] |
| Household contact (vs index case not in the same household) |  | 1.39 [0.95–2.04] |
| Children in household (yes vs no) |  | 1.49 [0.96–2.31] |
| **Outcome: More time to relax** |  |  |
| Age (per 10 years, reference age 18) |  | 0.98 [0.87–1.10] |
| Sex (male vs female) |  | 0.92 [0.63–1.34] |
| Working (possible or partially possible vs not possible) |  | 0.66 [0.44–0.98]* |
| Taking care of index case (yes vs no or not applicable) |  | 1.08 [0.69–1.69] |
| Children in household (yes vs no) |  | 0.59 [0.37–0.92]* |
| **Outcome: Change in depression score (during quarantine minus before)** |  |  |
| Age (per 10 years, reference age 18) |  | 1.02 [0.99–1.06] |
| Sex (male vs female) |  | 4.68 [1.67–13.12]* |
| Quarantine duration (per additional day) |  | 1.01 [0.81–1.27] |
| Living alone (vs living with others) |  | 0.91 [0.20–4.23] |
| Children in household (yes vs no) |  | 2.64 [0.75–9.31] |
| Working (possible or partially possible vs not possible) |  | 1.19 [0.38–3.64] |
| Baseline depression score (per additional point) |  | 1.23 [1.10–1.36]* |

### Difficulty of quarantine measures

Quarantine measures were perceived as difficult or very difficult by 84 participants (21.5%) during quarantine (Figure S3) and by 65 participants (17.5%) at the end of quarantine (Figure S4). In total, 109 participants (27.9%) found quarantine measures difficult or very difficult at either time point. Difficulty overall was lower for older adults (odds ratio 0.84 [95% CI: 0.75–0.95] for each additional 10 years), and lower for male participants (odds ratio 0.50 [95% CI: 0.34–0.72]) (Table 2). Although several participants commented on the difficulty of taking care of children and the infected person, we could not confirm with statistical significance that difficulty was higher for participants living with children (odds ratio 1.49 [95% CI: 0.96–2.31]), or for household contacts (odds ratio 1.39 [95% CI: 0.95–2.04]). There was no association between the overall difficulty and work from home during quarantine (odds ratio 1.00 [95% CI: 0.67–1.47]).

### Symptoms of depression, anxiety, and stress

370 participants answered the DASS-21 questionnaire both at baseline and at the end of quarantine. The proportion of participants with depressive symptoms (of any severity) doubled from 9.3% before quarantine to 18.9% during quarantine (Figure 4). A sensitivity analysis with all participants showed similar results (Figure S5). Overall, 65 participants (17.6%) had a relevant worsening by at least one category in depression, anxiety, or stress (Figure S6). However, 27 participants (7.3%) improved by at least one category in at least one subscale. Depression and stress scores increased with statistical significance (depression: +1.70 [95% CI: 1.19–2.22], stress +1.06 [95% CI: 0.47–1.66]) (Figure S6). The change in depression scores was higher in men than in women and higher in those with higher baseline scores, i.e., more pronounced in those with pre-existing depressive symptoms (Table 2).

**Figure 4:**
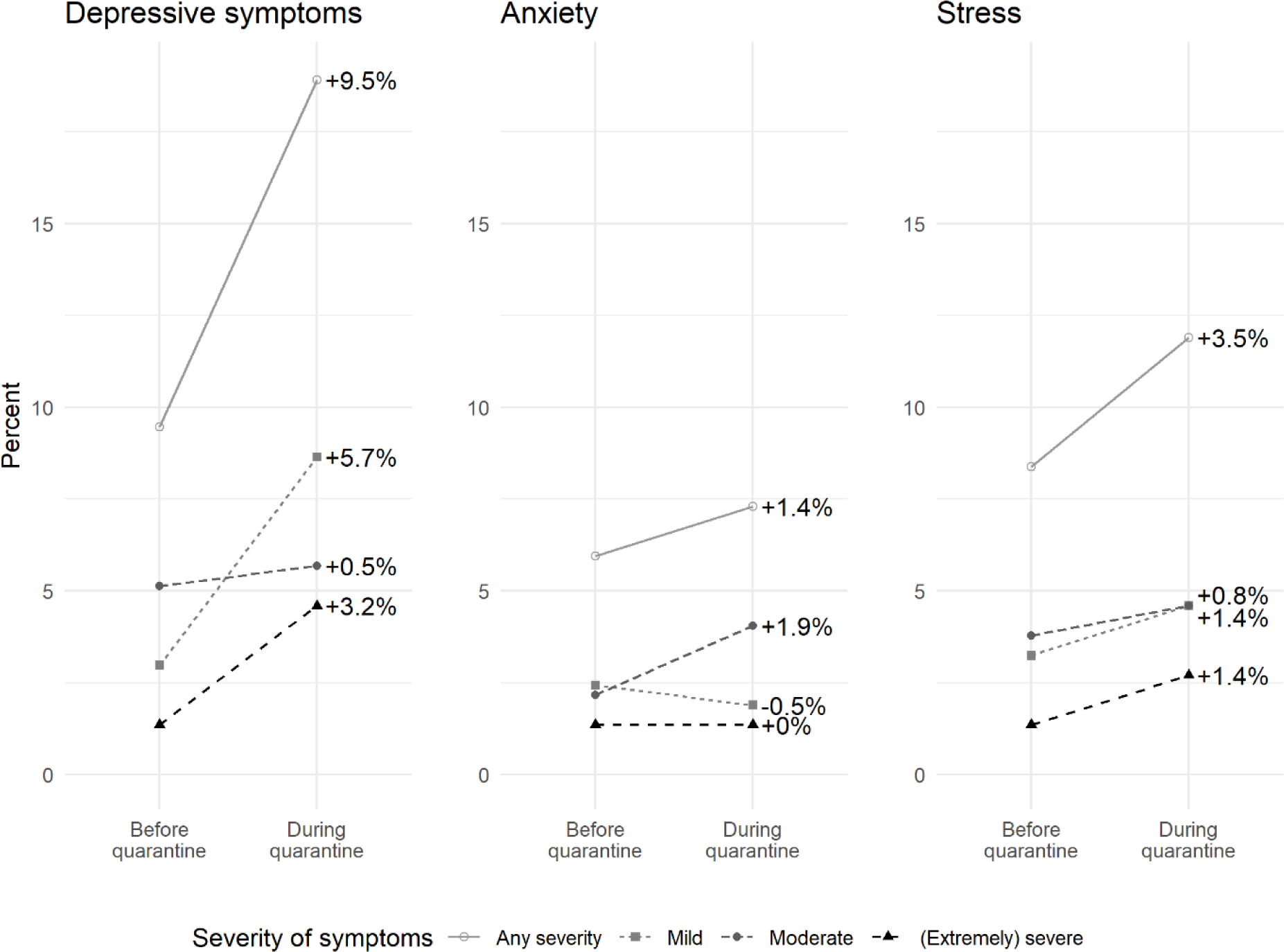
Change in proportion of persons with symptoms of depression, anxiety, or stress from before to during mandated quarantine. This analysis is restricted to participants who completed both questionnaires.

In direct comparisons, some participants reported feeling more isolated (22.0%) and increased trouble sleeping (14.0%) during quarantine compared to two weeks prior (Figure S7). Feeling more impatient or angry, consuming more alcohol, having more nightmares, feeling more worried, nervous, or depressed was reported by 8.3% to 11.0%.

### Positive experiences

Most participants reported more time to relax (253, 68.0%). Some commented that quarantine was stressful because they took care of children or of the SARS-CoV-2 positive person. An ordinal logistic regression confirmed that persons living with children (odds ratio 0.59 [95% CI: 0.37–0.92]) and persons working from home (odds ratio 0.66 [95% CI: 0.44–0.98]) had lower odds of reporting more time to relax.

### Information and preparation for quarantine

Most participants felt well or very well prepared for quarantine (47.2%) or neutral (40.3%). Older adults felt better prepared (odds ratio 1.19 [95% CI: 1.05–1.34] per additional 10 years), as did participants who felt well or very well informed (odds ratio 5.27 [95% CI: 3.03–9.32]). Participants worried about becoming depressed felt less well prepared (odds ratio 0.37 [95% CI: 0.23–0.58]).

### Adherence with quarantine

By the end of quarantine, 74 participants (19.9%) had left their house or met people during quarantine. One participant decided to stop quarantine early after a negative test based on the mistaken belief that a negative test ends quarantine.

We also asked household contacts about their adherence to the recommendations, in particular interacting with the person infected with SARS-CoV-2 (Figure S7). The recommendation that was least followed was wearing a mask when entering this person’s room – 20% rarely or never followed this rule.

Several participants expressed a need for physical activity outdoors and took precautions. One person described: “I badly needed a 20-minute walk or run. I went once early in the morning and once in the evening, so I virtually met nobody […]. I always had a mask with me. This should be allowed in my opinion.” Others answered that quarantine rules did not consider the need to walk the dog or helping people with disabilities.

### Financial hardship

In total, 62 of 390 participants worried about financial consequences: they were worried about losing their job or getting into financial difficulties or expected income loss due to quarantine (with no or only partial compensation). Only 14% (53 of 390) expected a reduced income; among them, 20 expected a partial, 25 no compensation. Twenty-two persons (6%) were worried, very, or extremely worried about getting into financial difficulties due to quarantine, and 39 persons (10%) were a little worried. 26 (7%) were a little worried about losing their job due to quarantine, and 12 persons were worried, very, or extremely worried.

### Burden of quarantine

In total, 176 of 390 participants (45.1%) found quarantine difficult, expected negative financial consequences or had symptoms of decreased mental health. There was little overlap between the 65 participants who worsened significantly in a DASS-21 dimension and the 62 participants who expected negative financial consequences; 117 of 390 participants either had worsened mental health or expected financial consequences (29.2%). Finally, 109 of 390 participants felt that quarantine measures were difficult or very difficult.

## Discussion

This study puts infection rates in close contacts (i.e., the secondary attack rate) into context with the negative and positive experiences of persons in quarantine. We found that about 11% of contacts were infected, and the majority (9%) had already converted by day 5 after the last (potentially infectious) contact. While some persons had positive experiences (e.g., more time to relax was reported by 68%), this stands in contrast to significant difficulties and burden experienced by almost half of the close contacts, including decreased mental health and financial consequences.

Our results compare to published secondary attack rates in close contacts, although these varied widely, partly due to different definitions of close contacts [29–34]. Even in household contacts, estimates ranged from 5% to 48% [29–34]. Highest estimates come from disadvantaged populations [34], reflecting that our population had comparatively good resources to protect themselves from transmission within a household.

Our study found a comparable increase in the risk for depressive symptoms as studies from earlier epidemics, which reported a 2.0-2.8 fold increase [4]. Prior studies also reported a higher risk among those with a history of depressive symptoms or mental illness, but did not report that the increase was higher in men [4]. Although others found associations of the duration of quarantine with negative psychological effects [3,36], we did not find a similar effect. A study in Germany found higher psychological distress among infected persons in isolation than in close contacts [37]. In comparison, our cohort study found similar rates of depressive symptoms for both groups [17]. However, in our study close contacts had higher rates of stress but lower rates of anxiety [17].

Although close contacts reported high motivation, adherence was moderate and comparable to other studies [1,5,6,38]. About 20% left their house, e.g., for groceries, or met persons from other households, and only half the household contacts reported voluntary self-quarantine before receiving the official mandate. In comparison, in the index case study arm of our cohort, 14% did not comply with measures before their positive result, and 3% after their positive result [17]. This is in line with other studies where adherence was lower in persons without symptoms [38]. We found that contacts who worried less about contracting the virus were less motivated to adhere. Contact tracers should therefore carefully explain why the exposure was potentially infectious.

Quarantine should be limited to the shortest possible but necessary duration. A 10-11 day quarantine was sufficient in our population, in a setting with high testing resources. Our study also adds to other evidence that supported a shortened 7-day quarantine with a negative test at the end of quarantine, as introduced soon after our study concluded [12–14,39]. Public health authorities should provide better information on testing. For example, 7% of our participants experienced long quarantine durations of 12 to 20 days due to others in their household testing positive. These extensions could have been reduced if their household members had been tested earlier.

Future interventions should aim to reduce the burden of quarantine for parents and persons with depressive symptoms [3]. Participants in our survey expressed a strong desire for outdoor physical activity, walking their dog, and letting children go outside. Experiences among parents are in line with our findings in infected persons in isolation, where parents had higher difficulty with isolation measures than persons without children [17].

A limitation of our study is that we only included close contacts of adult index cases, although sometimes children in the same household were also positive. Parents likely experienced more difficult quarantines, so we may have underestimated difficulties. Furthermore, we may have overestimated motivation and adherence as more motivated persons may have been more likely to consent to participate in the study. Additionally, we did not measure long term effects on mental health, although a prior study reported sustained negative effects 6 months after quarantine [3]. Finally, infection rates among close contacts alone are not sufficient to measure quarantine’s effectiveness, which also depends on epidemic severity, timeliness of contact tracing, duration and adherence with quarantine [40]. Delays in contact tracing and many undetected or not traced COVID-19 cases during the Fall 2020 surge likely limited quarantine’s effectiveness, but we did not aim to estimate averted infections.

To inform future pandemic response and evaluate past public health interventions, it is important to compare the effectiveness with the burden placed on quarantined persons. Our analysis exemplifies that this comparison should ideally be performed in the same population, as many factors, such as mental health, motivation, and adherence, influence each other. Going forward, policy makers should critically assess the need for quarantine and strategically study and reduce negative impact on quarantined persons.

## Data Availability

We are open to sharing individual participant data that underlie the results reported in this article, after de-identification upon reasonable requests to the corresponding author. Data requestors will need to sign a data access agreement.

## Acknowledgments

We thank Prof. Torsten Hothorn for his support with the survival analysis.

## Patient consent statement

All participants provided electronic consent. The study was prospectively registered (ISRCTN14990068) and approved by the ethics committee of the Canton of Zurich (BASEC 2020-01739).

