## Supplement for "SARS-CoV-2 quarantine mandated by contact tracing: burden and infection rate among close contacts in Zurich, Switzerland, 2020-2021"

#### Participants vs non-participants

*Table S1: Comparison of participants and non-participants based on data from contact tracing only. *Duration of quarantine was estimated using the actual recorded date of first contact with the close contact (email, phone call or text message) from contact tracing as the start date of quarantine (pessimistic estimate). In a more optimistic estimate, based on the appeal from the Department of Health to index cases to inform their close contact directly, uses the date of the index cases' positive test result as the start date of close contacts' quarantine.*

| | | **Participants** | | **Non-participants** | |
|---|---|---|---|---|---|
| | | **N of 395** | **%** | **N of 1018** | **%** |
| Age at report date (years) | 18-39 | 187 | **47** | 395 | **39** |
| | 40-64 | 130 | **33** | 180 | **18** |
| | 65+ | 25 | **6** | 81 | **8** |
| | missing | 53 | **13** | 362 | **36** |
| Gender | Female | 190 | **48** | 512 | **50** |
| | Male | 194 | **49** | 453 | **44** |
| Age of index case at report date (years) | 18-39 | 229 | **58** | 454 | **45** |
| | 40-64 | 113 | **29** | 313 | **31** |
| | 65+ | 53 | **13** | 251 | **25** |
| Duration of quarantine (days)* | | **Mean** | **SD** | **Mean** | **SD** |
| | Pessimistic estimate | 7.9 | 1.9 | 7.5 | 2.1 |
| | Optimistic estimate | 9.1 | 1.6 | 8.7 | 1.6 |

*Table S2: Mean and standard deviation of close contacts' age by age group of the index case, for participants and non-participants. Younger index cases had younger close contacts on average.*

| | **Age of close contacts: mean (SD)** | |
|---|---|---|
| **Age range index cases** | Participants | Non-participants |
| All ages (18+) | 41 (15) | 40 (17) |
| Age 18-39 | 35 (12) | 31 (11) |
| Age 40-64 | 47 (14) | 42 (15) |
| Age 65+ | 62 (12) | 59 (18) |

### Efficacy of quarantine: positive PCR tests during quarantine

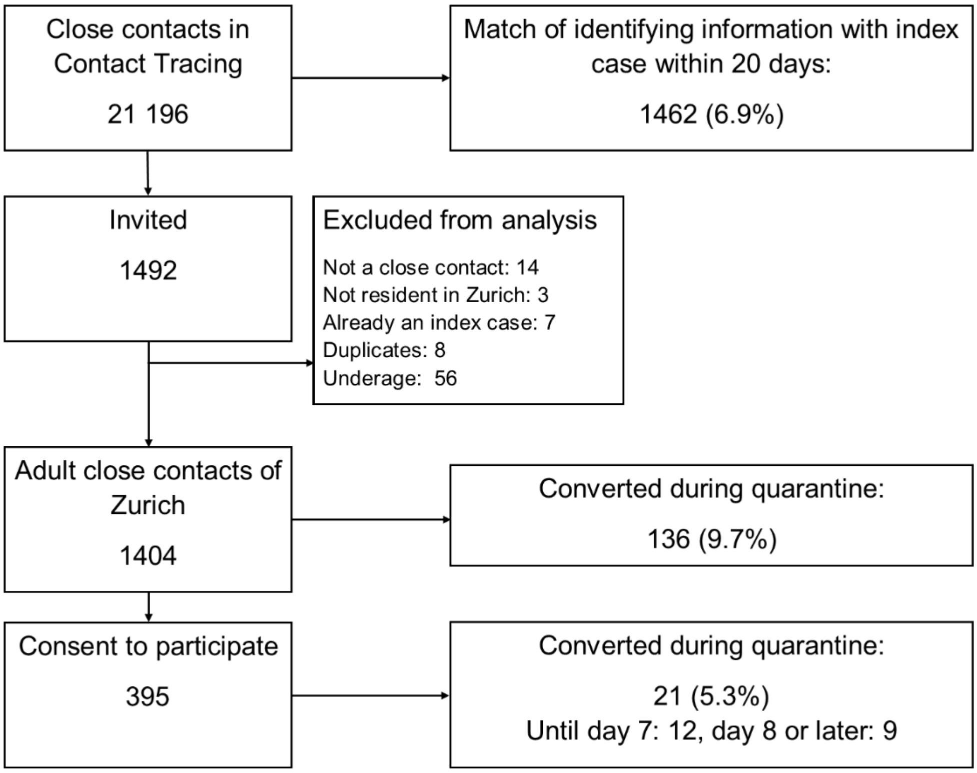


*Figure S 1: Converted persons in the study flow. Based on matches of name and birthdate, 6.9% of all close contacts (including children) converted within 20 days. Here, we did not consider persons who were not close contacts in the denominator. However, birthdates were often missing and names often misspelt. Among adult close contacts of Zurich, we found 9.7%. This information is based on matches of name and birthdate, but also information from close contacts during the recruitment process. Those who had already converted before entering the study were invited as index case instead. Therefore, only 5.3% of close contacts in the study converted within 20 days.*

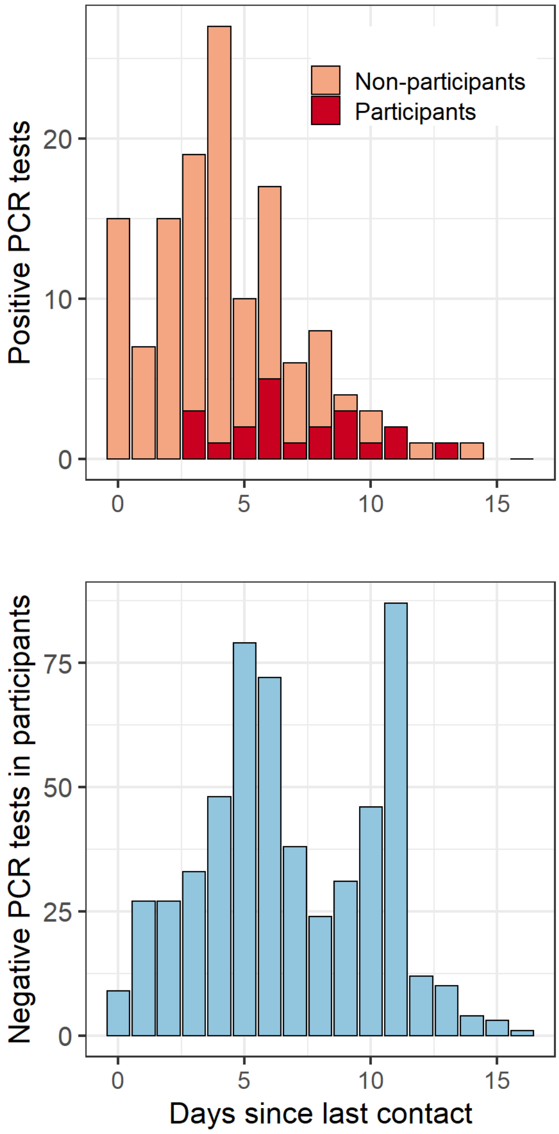


*Figure S 2: A) Positive tests in participants and non-participants. Participants were positive later because if they converted earlier, they were invited to a different study arm, and count as non-participants here.*
*B) Negative PCR tests during quarantine. Repeated negative tests are possible within individuals.*

#### Difficulties during quarantine

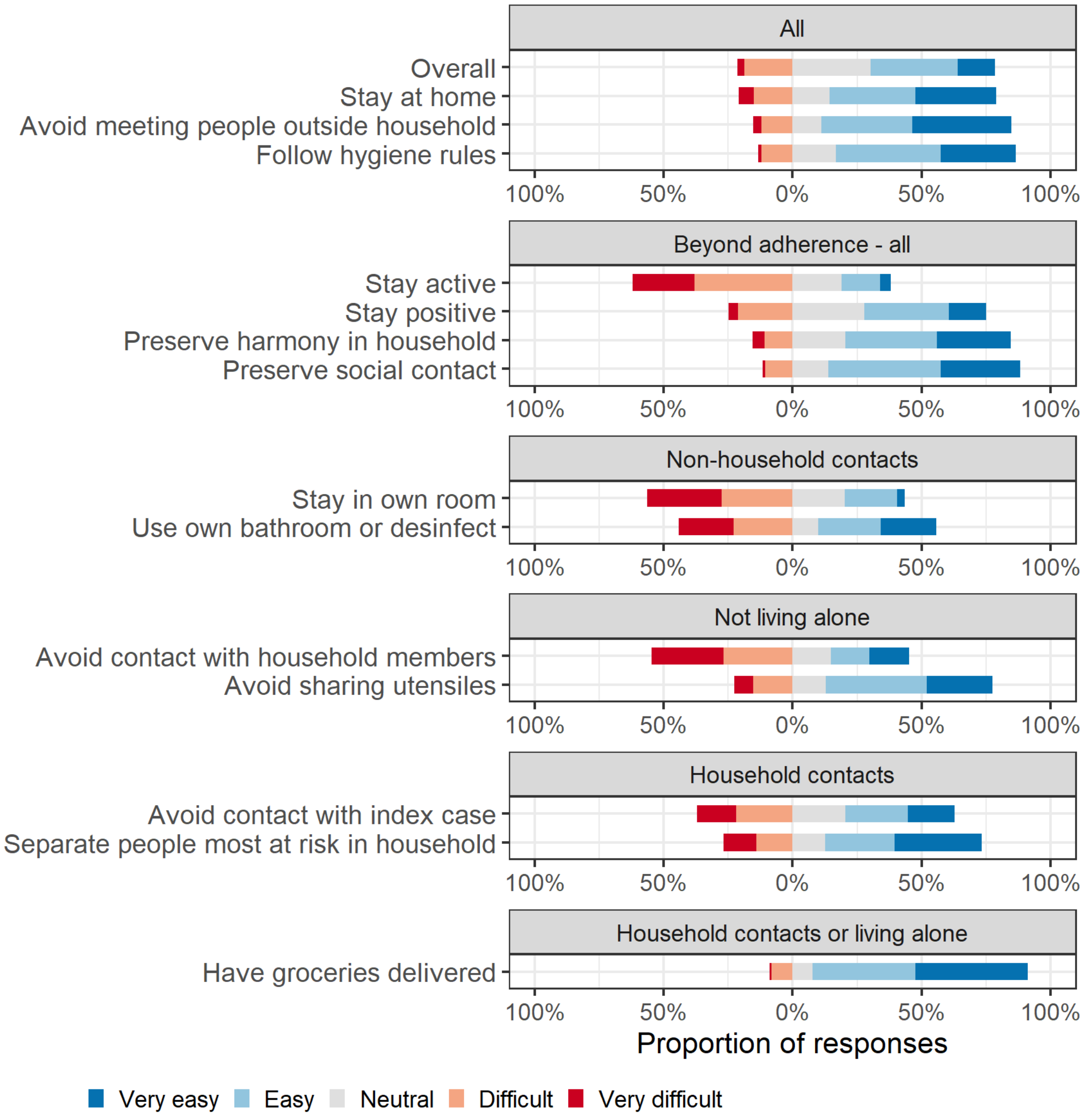


*Figure S 3: Difficulties during quarantine*

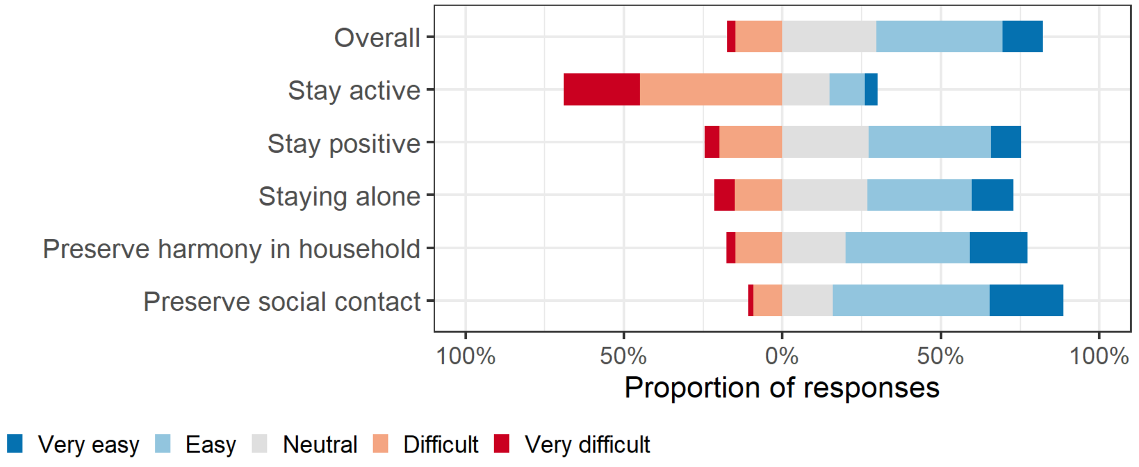


*Figure S 4: Difficulties re-evaluated at the end of quarantine*

#### Mental health

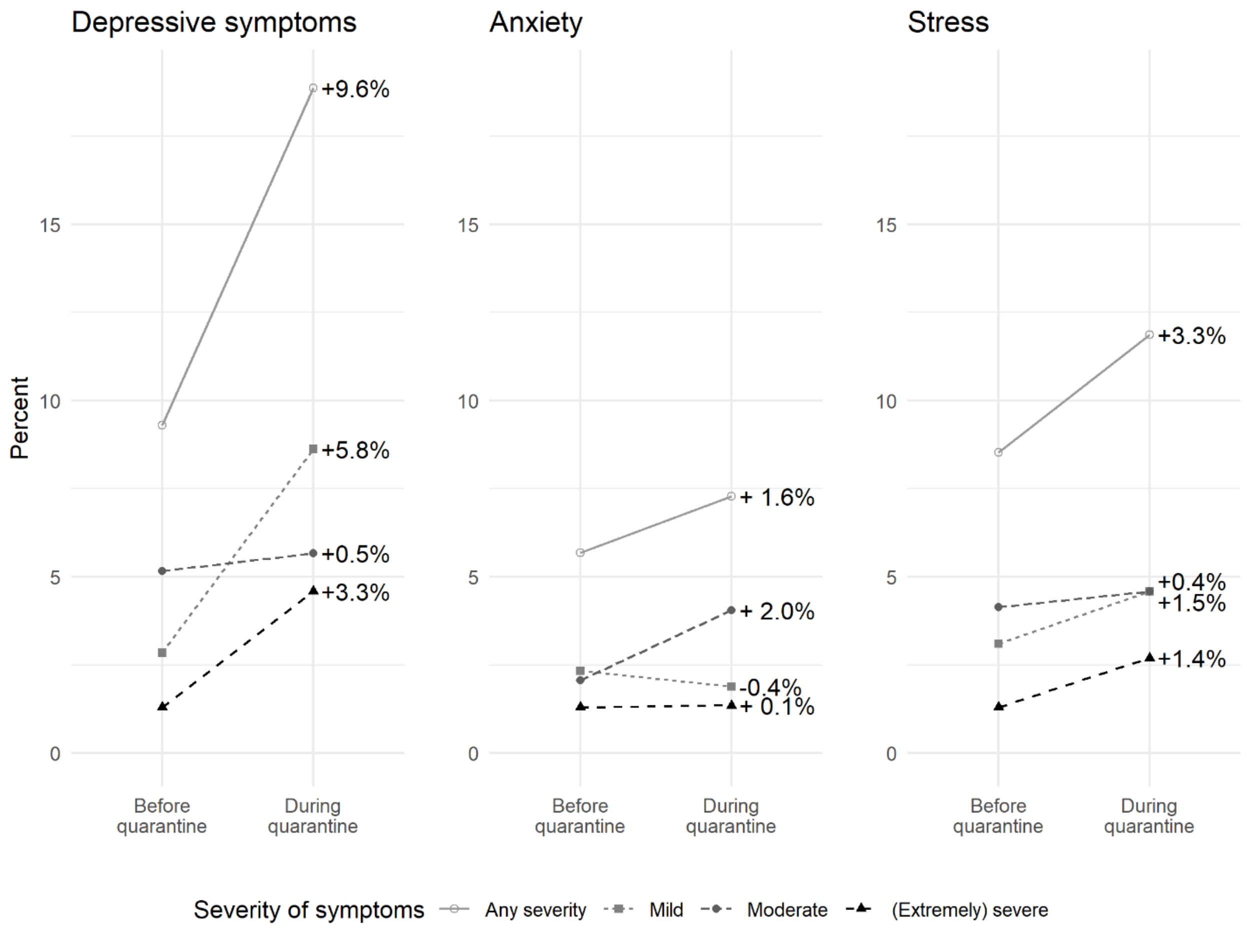


*Figure S5: Change in DASS categories among all respondents (387 at baseline and 371 at end of quarantine), not restricted to those who answered both questionnaires.*

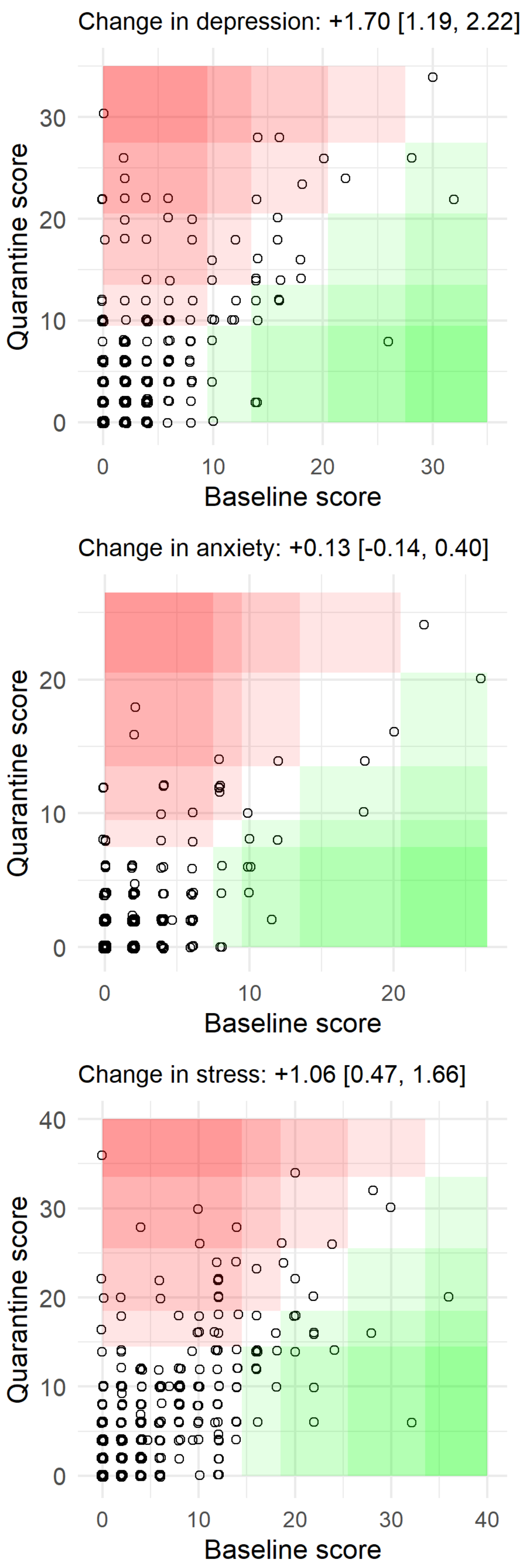


*Figure S6: Change in symptoms of depression, anxiety and stress between baseline and during quarantine on the DASS scale. Shaded areas indicate the thresholds of symptom categories (normal, mild, moderate, severe, extremely severe). Participants in red areas experienced a worsened state by at least one category (darker shaded areas indicate a jump in more categories). Participants in green areas improved during quarantine. The average change in scores is indicated as mean with a 95% confidence interval based on the standard error.*

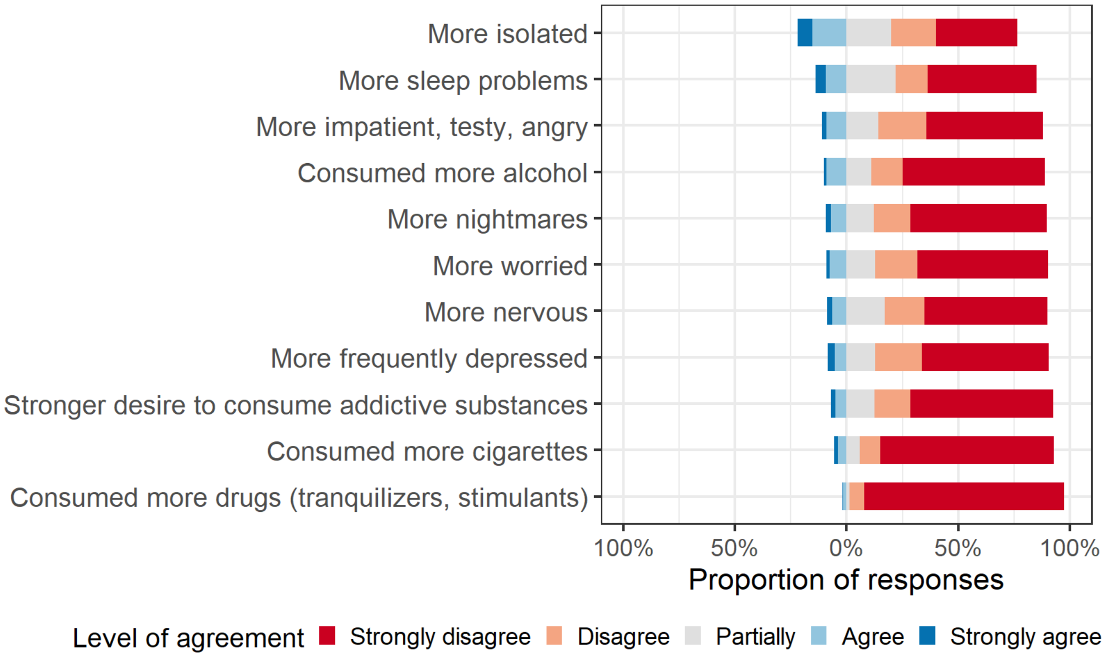


*Figure S 7: Comparison of mental health during and before quarantine*

#### Financial hardship

When asked during quarantine whether they expect to experience a reduced income due to the quarantine, 76% (296 of 390) answered no. 14% (53 of 390) answered yes, among them 8 expected a complete compensation, 20 a partial compensation, and 25 did not expect any compensation. 19 answered maybe and 22 did not know or preferred not to answer.

Of the 45 persons who expected a reduced income without compensation or only partial compensation, 21 were employed, 15 were self-employed, 2 were both self-employed and employed, 5 were students and 2 were retired. Most participants in our study were employed (273 or 70%) and only 24 or 6% were self-employed or both self-employed and employed. Thus, 17 of 24 (71%) self-employed persons expected a reduced income. In comparison, 217 of 273 (79%) employed participants did not expect a reduced income.

22 persons (6%) were worried, very worried or extremely worried about getting into financial difficulties due to quarantine. Oddly, 8 of them did not expect a reduced income, and 2 did but expected full compensation. Among the 22 persons worried about financial difficulties, only 1 was self-employed. 39 persons (10%) were a little worried, and 82% (320 of 390) were not at all worried about financial difficulties due to quarantine.

87% (338 of 390) of participants were not at all worried about loosing their job due to quarantine, and 26 (7%) were only a little worried. Among 12 persons worried, very worried or extremely worried about loosing their job due to quarantine, 9 answered that they were employed, 1 was a student, 1 person was retired, and oddly, one answered that they were unemployed.

#### Adherence of household contacts and their index cases to quarantine measures

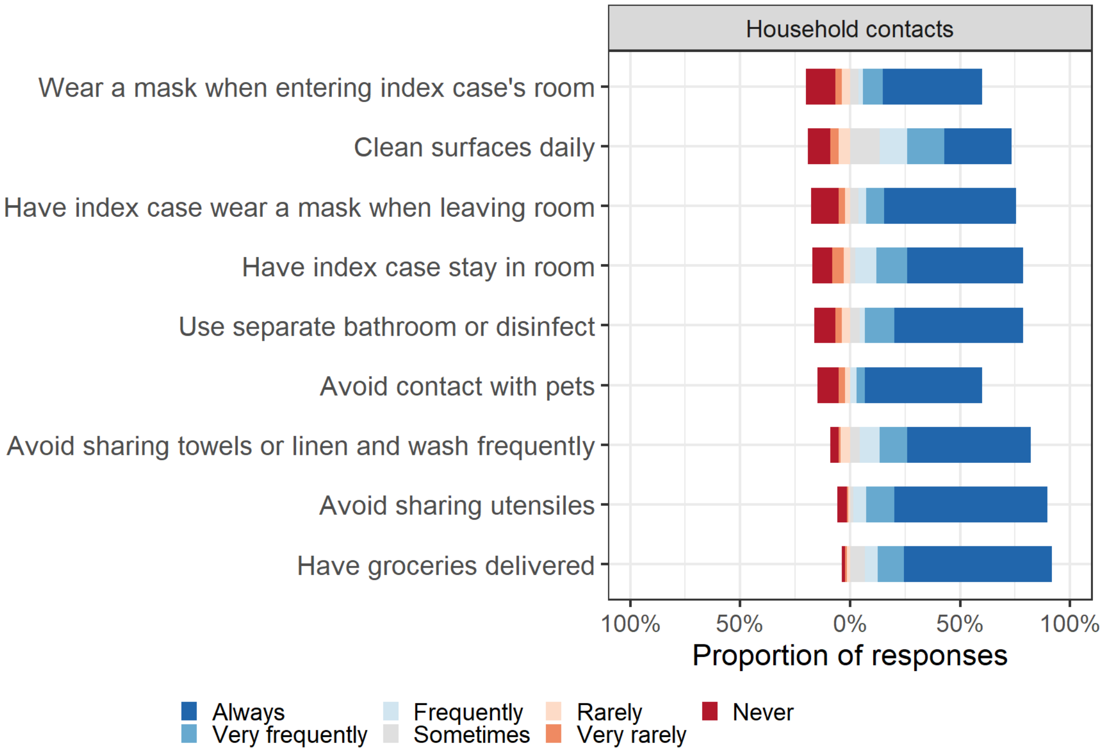


*Figure S 8: Adherence of household contacts and their index cases to quarantine and isolation measures*

##### Close contacts leaving their house

197 respondents (53%) reported having left the house for one or more reasons. The most frequent reason was to get a SARS-CoV-2 test (158 respondents), followed by physical activity (20 respondents), buying groceries (14 respondents), need for health care (9 respondents), walking the dog (8 respondents), to help or meet family or friends (6 respondents), switch of quarantine place (3 respondents), to take out trash, get mail, or for gardening (6 respondents), to work (2 respondents), for mental health (2 respondents).

##### Close contacts meeting persons

Most respondents did not meet anyone outside their household during quarantine (319 respondents or 85.8%), and 7 respondents only met a health care provider or our study team. Forty respondents were potentially not compliant: they declared having met family (21), friends (15), neighbors (5), deliverymen (3), co-workers (3) or others (3). A few respondents specified in comments that they met these people while keeping distance (2), from their balcony (2). Two offered supposedly extenuating circumstances: one met a friend for coffee who had recovered from SARS-CoV-2, and one met a friend who lived alone.

Only 7 respondents both met someone outside their household (other than a health care provider or the study team) and left the house. Conversely, up to 74 respondents (19.9%) admitted not fully complying with quarantine measures with respect to not meeting anyone and not leaving their house.

#### Background on contact tracing in the canton of Zurich, Switzerland

The cantonal Department of Health recommended a test on day 5, regardless of symptoms, at the beginning of the study, and but dropped this recommendation end of October 2020, and recommended instead to always test upon symptom onset. However, the federal office of public health (FOPH) did not change their recommendation to test in the beginning of quarantine, and the test remained free of charge for all close contacts during quarantine.

The ZSAC study was closely linked to contact tracing and adapted to its dynamic changes. Therefore, the most relevant changes in contact tracing over time are briefly described here. Contact tracing in Zurich officially started on May 11, 2020, and was largely performed through telephone calls by a team of security staff from the Zurich airport deployed for this purpose, under the medical supervision of the Department of Health, and later of JDMT, a private company. After contact tracing was overwhelmed by the rising number of cases in early October 2020, more automated processes were introduced. By mid-October, only index cases were called anymore (with great delays), and they were instructed to inform their close contacts themselves. Close contacts received a text message from contact tracing. By November 2020, an online form was launched, where index cases could complete a basic questionnaire and declare their close contacts. This online form triggered a text message to the close contact, giving access to another online form for the close contact to download the official document of the mandated quarantine. In parallel, index cases were still called by phone.
